# Prediction Modeling of Missed Appointments in Safety-Net Physiatry: Visit Characteristics Outperform Patient Risk Scores

**DOI:** 10.1101/2025.04.26.25326474

**Authors:** Hai Trinh, Robertus Kounang

**Affiliations:** Department of Physical Medicine and Rehabilitation, Arrowhead Regional Medical Center, Colton, CA, USA

**Keywords:** missed appointments, no-show, physical medicine and rehabilitation, safety-net hospital, social determinants of health, telehealth, prediction model, electronic health records

## Abstract

**Objective:** To identify appointment-level and patient-level factors associated with missed (no-show) visits in an outpatient physical medicine and rehabilitation (PM&R) clinic at a public safety-net hospital, to test whether electronic health record (EHR) composite risk scores and neighborhood social vulnerability add predictive value beyond scheduling characteristics, and to develop and internally validate a no-show prediction model from routinely available scheduling data.

**Design:** Retrospective cohort study of all scheduled outpatient PM&R appointments, with development and internal (cross-validated and temporal) validation of a prediction model.

**Setting:** Outpatient PM&R clinic, Arrowhead Regional Medical Center, a county safety-net hospital in the Inland Empire region of Southern California.

**Participant:** 8,927 appointments scheduled for 2,409 adults between February 15, 2022 and February 12, 2026; 6,594 appointments for 1,369 patients were linked to age, EPIC General Risk Score, EPIC Preventive Care Gap Score, and ZIP-code Social Vulnerability Index (SVI).

**Main outcome measure:** No-show at the appointment, defined as a scheduled appointment neither attended nor cancelled; cancellations were retained as a separate status.

**Results:** The no-show rate was 13.1% (1,171 of 8,927); 29.0% of appointments were cancelled. Ten percent of patients accounted for 50% of no-shows. In generalized estimating equation models clustered by patient, no-shows were more likely at new-patient consultations than follow-ups (OR 1.40, 95% CI 1.10-1.78) and less likely at telehealth visits (OR 0.25, 95% CI 0.14-0.44), procedure visits (OR 0.27, 95% CI 0.09-0.77), same-day appointments (OR 0.59, 95% CI 0.35-1.01), and with increasing age (OR 0.88, 95% CI 0.81-0.96 per decade). The EPIC General Risk Score (OR 0.99, 95% CI 0.94-1.04), Preventive Care Gap Score (OR 1.02, 95% CI 0.96-1.09), and both SVI measures were not associated with no-show. A logistic model using only scheduling features discriminated no-shows with a patient-grouped cross-validated area under the ROC curve (AUC) of 0.660 (95% CI 0.636-0.681); adding age, EPIC scores, and SVI did not improve discrimination (AUC 0.679 vs 0.679 in the linked cohort). Temporal validation on 2025-2026 appointments gave an AUC of 0.624 (95% CI 0.593-0.658). Sex, race/ethnicity, and completion of SDOH screening were not associated with no-show in the fully linked subset.

**Conclusions:** In safety-net PM&R, missed visits are associated with how a visit is scheduled rather than with EHR-derived clinical risk, neighborhood vulnerability, or demographic characteristics. Automatically generated composite risk scores should not be assumed to identify patients at risk of non-attendance. Scheduling-level levers, in particular telehealth and targeted outreach before new-patient consultations, are the more promising focus for reducing no-shows in this setting.

## Introduction

Missed outpatient appointments delay care, waste clinical capacity, and disproportionately affect patients who already face barriers to access. In physical medicine and rehabilitation (PM&R), where functional gains depend on sustained follow-up, therapy, and procedural care, the clinical cost of non-attendance is particularly high.1-3 Public safety-net hospitals rarely discharge patients for non-attendance, so a subset of patients with repeated missed visits can absorb a large share of scheduling capacity.

A large literature has modeled no-show risk, most often in primary care, radiology, and specialty clinics, and generally finds that scheduling characteristics-lead time, visit type, prior attendance, and modality-are the strongest predictors, with discrimination in the range of an area under the receiver operating characteristic curve (AUC) of 0.70 to 0.85.4-7,15 Whether the composite risk metrics that modern EHR platforms generate automatically, such as EPIC’s General Risk Score and Preventive Care Gap Score, carry additional information about attendance behavior has received little attention. These scores are attractive because they are already displayed for every patient and require no additional data collection. There is similar interest in whether area-level social vulnerability indices or individual social drivers of health (SDOH) screening improve prediction or identify structural barriers.4,6,10,11,14 PM&R populations in safety-net settings, who are often older, functionally limited, and dependent on public insurance and informal caregivers, are under-represented in this literature.

The Inland Empire region of Southern California is a relevant setting. It is heavily car-dependent with limited public transit, has a high cost of living relative to income, and serves a population that is majority Hispanic/Latino and predominantly insured through Medi-Cal, Medicaid, or Workers’ Compensation.8,12 ZIP-code social vulnerability in the clinic’s catchment area is among the highest in California. These characteristics are commonly assumed to drive non-attendance, yet whether they are visible in structured EHR data is unknown.

An earlier analysis of this clinic’s patients, posted as a preprint, used patient-level counts of missed appointments and reported that EPIC composite scores were strongly associated with the number of missed visits. That analysis had two limitations that motivated the present study: it lacked a denominator of scheduled appointments, so it could not distinguish patients who missed many appointments because they had many scheduled from patients who missed a high proportion of them; and its missed-appointment count was drawn from a hospital-wide field rather than from PM&R scheduling records. The present study uses appointment-level PM&R scheduling data to address both.

Our aims were: (1) to describe the distribution of no-shows across appointments and patients in a safety-net PM&R clinic; (2) to estimate the independent associations of appointment characteristics, patient age, EPIC composite scores, and neighborhood social vulnerability with no-show at the appointment level, accounting for clustering within patients; (3) to develop and internally validate a prediction model, intended for use by clinic scheduling staff, that identifies appointments at elevated risk of no-show from routinely available scheduling data; and (4) to test whether EHR composite scores and social vulnerability improve that model. As a secondary methodological aim, we compare patient-level and appointment-level analyses of the same patients to show how the choice of analytic unit changes conclusions.

## Methods

### Study design and setting

This was a retrospective cohort study of scheduling data from the outpatient PM&R clinic at Arrowhead Regional Medical Center (ARMC), a public county hospital in Colton, California.

The clinic operates three days per week (Tuesday through Thursday) and sees approximately 12 to 18 patients per clinic day. Data were extracted from the Epic EHR for all outpatient PM&R appointments scheduled between February 15, 2022, the earliest date available in the institution’s Epic scheduling records, and February 12, 2026. The study is reported in accordance with the TRIPOD+AI statement for prediction-model studies16 and the STROBE statement for observational studies.17 No protocol was registered.

### Data sources and linkage

Three de-identified extracts were used. The scheduling extract, obtained from Epic Clarity by the institution’s research data office, contained one row per scheduled appointment with appointment date, visit type, appointment length, telehealth flag, department, appointment status, cancellation reason and date, and same-day and walk-in flags. Two earlier patient-level extracts from Epic SlicerDicer supplied, for a subset of patients, age, EPIC General Risk Score, EPIC Preventive Care Gap Score, and 2020 CDC/ATSDR Social Vulnerability Index (SVI) percentiles for ZIP code (the risk-score extract), and legal sex, race/ethnicity, and SDOH screening results (the demographic extract). Extracts were linked by a study-specific pseudonymous patient identifier assigned by the data office; no direct identifiers were retained. SlicerDicer emits one row per value for multi-valued fields, so the patient-level extracts were collapsed to one row per patient before linkage.

Of 2,409 patients in the scheduling extract, 1,369 (56.8%) were present in the risk-score extract and 887 (36.8%) in both patient-level extracts (Figure 1). Patients absent from the patient-level extracts were more often new to the clinic (44.6% of their appointments were first visits vs 20.8%), had fewer appointments (mean 2.2 vs 4.8), and had a higher no-show rate (18.3% vs 11.3%). Analyses were therefore structured in three nested cohorts: the full cohort (all appointments; description and scheduling-feature models), the linked cohort (appointments for patients with age, EPIC scores, and SVI; primary adjusted analysis), and the fully linked cohort (additionally with sex, race/ethnicity, and SDOH screening; sensitivity analysis). Analysis was complete-case within each cohort; no predictor had missing values among linked patients.

**Figure 1.**
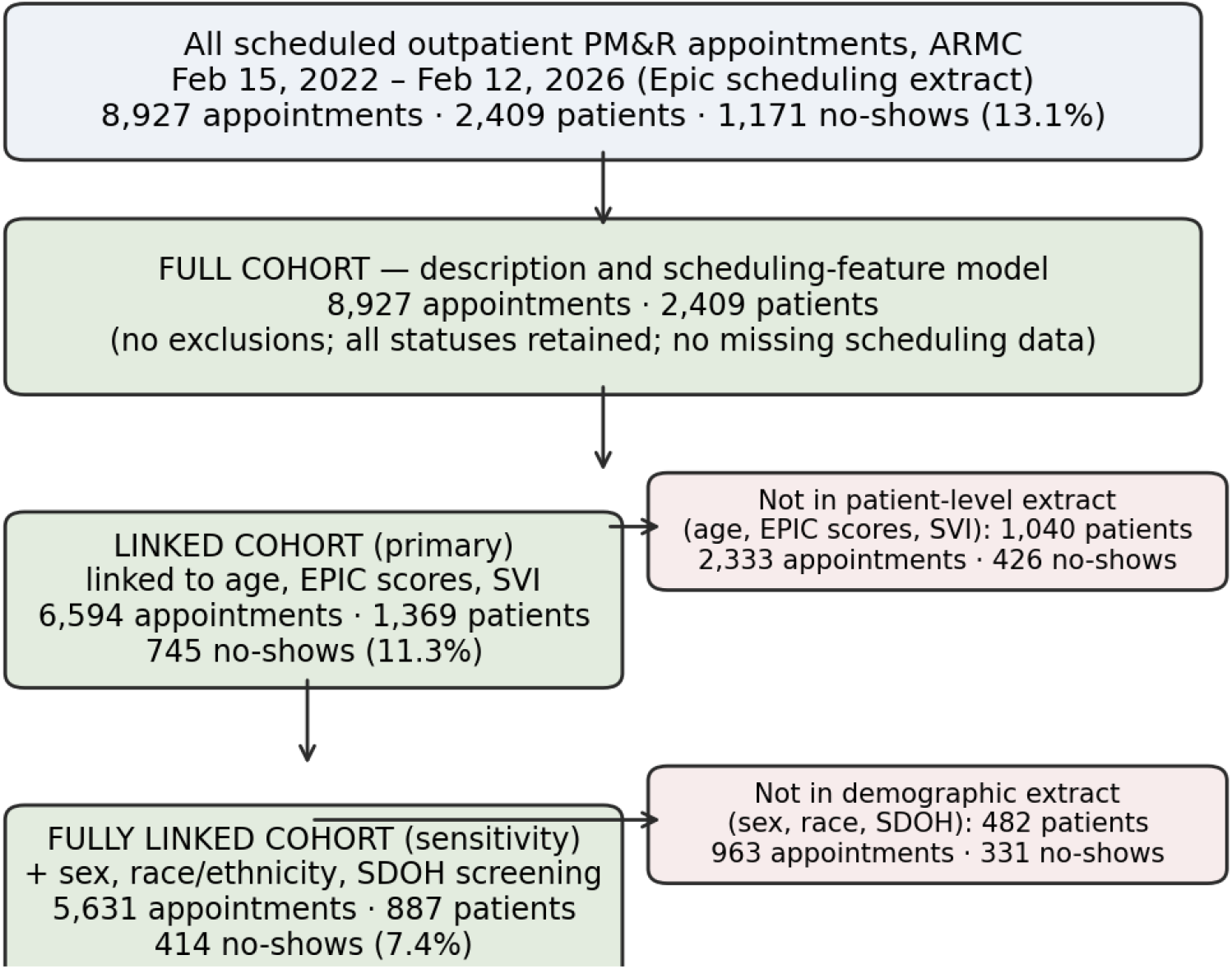
Flow of appointments and patients through the study.

### Participants

All scheduled outpatient PM&R appointments in the study window were included, regardless of status. No appointment-level exclusions were applied. Patient-level extracts had been restricted to adults aged 30 years or older; consequently, the linked cohorts comprise patients aged 30 or older, whereas the full cohort has no age restriction.

### Outcome

The outcome was no-show at the appointment, defined as a scheduled appointment recorded by scheduling staff as “No Show”: the patient neither attended nor cancelled. Appointments recorded as “Canceled” (by patient, provider, or system) were retained as a separate status and were not counted as no-shows; “Completed” appointments were the reference. The status was recorded at the time of the appointment, independently of and before any predictor was extracted, by the same procedure for all patients. No predictor was used to define the outcome.

### Predictors

**Appointment-level (scheduling) predictors, available for all appointments:** visit type (follow-up [reference], consultation, evaluation, procedure, or other); telehealth versus in-person; same-day appointment; day of week; calendar year; whether the appointment was the patient’s first in the clinic during the study window; and the patient’s number of prior no-shows and prior cancellations in the clinic before that appointment, computed strictly from earlier-dated appointments. Appointment length was collinear with visit type and was not modeled. Walk-in status was recorded for no appointment and was dropped. The date on which each appointment was booked was not available, so lead time could not be derived. **Patient-level predictors, available for the linked cohort:** age in years; EPIC General Risk Score, a composite of chronic conditions, prior hospitalizations, vital signs, and laboratory values; EPIC Preventive Care Gap Score, a count of overdue preventive services; and SVI percentiles for the patient’s ZIP code in the socioeconomic and housing/transportation themes. The minority-status SVI theme was omitted because of its near-ceiling distribution. **Additional predictors in the fully linked cohort:** legal sex; race/ethnicity as recorded in the EHR (Hispanic/Latino [reference], non-Hispanic White, non-Hispanic Black, Asian American/Native Hawaiian/Pacific Islander, unknown); and whether any SDOH screening result was recorded in any of 12 domains.

All predictors were prespecified as the full set available in the extracts; none was selected or removed on the basis of its association with the outcome. Patient-level values were recorded at the time of extraction rather than at each appointment (see Limitations).

### Statistical analysis

No-show rates were described overall and by appointment and patient characteristics. Because appointments are clustered within patients, adjusted associations were estimated with generalized estimating equations (GEE) for a binary outcome with a logit link, patient as the cluster, an exchangeable working correlation, and robust (sandwich) standard errors; results are odds ratios (ORs) with 95% confidence intervals. Three GEE models were fitted: scheduling predictors only, in the full cohort; scheduling plus patient-level predictors, in the linked cohort (primary); and the primary model plus sex, race/ethnicity, and SDOH screening, in the fully linked cohort. Continuous predictors were entered untransformed; age and SVI were scaled per 10 units. Multicollinearity was assessed with variance inflation factors.

Sample size. All eligible appointments were used. With 1,171 events in the full cohort and 745 in the linked cohort for at most 18 candidate parameters, the data provide approximately 90 and 41 events per parameter respectively, well above conventional minimums.18

Prediction model. The primary prediction model was a logistic regression using scheduling predictors only, fitted in the full cohort, because these are the features a scheduler can see for every appointment. A gradient-boosted tree classifier with default hyperparameters was fitted for comparison. Two internal validation strategies were used. First, 5-fold cross-validation stratified by outcome and grouped by patient-so that all of a patient’s appointments fell in the same fold and no patient contributed to both training and test data-repeated five times with different partitions. Second, temporal validation: the model was fitted to appointments from 2022 through 2024 and evaluated on appointments from 2025 through February 2026. Discrimination was summarized as AUC with 95% confidence intervals from 500 patient-level cluster bootstrap resamples of out-of-fold predictions; calibration as the calibration slope, calibration-in-the-large, Brier score, and scaled Brier score, with a calibration plot;19 and clinical yield as the proportion of no-shows captured by the highest-risk 20% of appointments. To test whether patient-level information adds value, the same models were refitted in the linked cohort with and without age, EPIC scores, and SVI. Discrimination was also estimated within subgroups defined by age, General Risk Score, sex, and race/ethnicity. No class-imbalance adjustment, hyperparameter tuning, or model selection on test-fold performance was performed. The model output is a probability; a risk-decile table is provided so that clinics can choose an outreach threshold matched to capacity.

Patient-level versus appointment-level comparison. For patients in the linked cohort, we aggregated PM&R appointments to one record per patient and fitted negative binomial models of (a) the number of no-shows, (b) the number of scheduled appointments, and (c) the number of no-shows with the log of scheduled appointments as an offset, each on age, EPIC scores, and SVI, and compared the resulting incidence rate ratios with the appointment-level GEE odds ratios for the same predictors.

Sensitivity analyses of the primary GEE model used an independence working correlation; excluded telehealth visits; excluded same-day appointments; excluded 2026 (partial year); and excluded patients with more than 20 appointments. Analyses used Python 3.12.3 with pandas 3.0.2, statsmodels 0.15.0, and scikit-learn 1.8.0; two-sided p < 0.05 was considered significant. Full code is provided as Supplementary File 1.

## Results

### Appointments and patients

The full cohort comprised 8,927 appointments for 2,409 patients (median 2 appointments per patient, range 1-85). Of these, 5,164 (57.8%) were completed, 2,592 (29.0%) were cancelled, and 1,171 (13.1%) were no-shows. Most cancellations were initiated by the patient (1,681) or provider (447); 68 were attributed to missing insurance authorization and 18 to financial reasons. Follow-up visits (56.0%) and consultations (39.5%) accounted for nearly all appointments; 129 (1.4%) were telehealth and 274 (3.1%) were same-day. 831 patients (34.5%) had at least one no-show, and the 10% of patients with the most no-shows accounted for 50% of all no-shows. Characteristics of the cohorts are shown in Table 1.

**Table 1.** Characteristics of appointments and patients in the three analytic cohorts.

| Characteristic | Full cohort | Linked cohort | Fully linked cohort |
| --- | --- | --- | --- |
| Appointments, n | 8,927 | 6,594 | 5,631 |
| Patients, n | 2,409 | 1,369 | 887 |
| Appointments per patient, median (range) | 2 (1-85) | 2 (1-85) | 2 (1-85) |
| No-shows, n (%) | 1,171 (13.1) | 745 (11.3) | 414 (7.4) |
| Cancellations, n (%) | 2,592 (29.0) | 1863 (28.3) | 1527 (27.1) |
| **Appointment characteristics, n (%)** |  |  |  |
| Follow-up visit | 5,002 (56.0) | 4,347 (65.9) | 4,134 (73.4) |
| Consultation | 3,529 (39.5) | 2,049 (31.1) | 1,319 (23.4) |
| Procedure | 130 (1.5) | 99 (1.5) | 91 (1.6) |
| Telehealth | 129 (1.4) | 125 (1.9) | 124 (2.2) |
| Same-day | 274 (3.1) | - | - |
| First visit in clinic | 2,409 (27.0) | 1,369 (20.8) | 887 (15.8) |
| **Patient characteristics, mean** |  |  |  |
| Age, years | - | 55.0 | 55.6 |
| General Risk Score | - | 2.17 | 2.32 |
| Preventive Care Gap Score | - | 4.37 | 4.17 |

Full cohort: all scheduled PM&R appointments. Linked cohort: appointments for patients with age, EPIC scores, and SVI available. Fully linked cohort: additionally with sex, race/ethnicity, and SDOH screening. Same-day flag was available only in the scheduling extract and is shown for the full cohort. Patient-level extracts were restricted to age $\geq 30$ ; the full cohort has no age restriction.

Unadjusted no-show rates varied substantially by appointment characteristics (Figure 2). Consultations had a no-show rate of 18.8% compared with 9.6% for follow-ups and 3.8% for procedures. Telehealth visits had a rate of 2.3% versus 13.3% in person. A patient’s first visit in the clinic had a rate of 17.9% versus 11.4% for subsequent visits. The rate rose from 9.6% in 2022 to 16.2% in 2024 before falling to 11.9% in 2025 (Figure 6).

**Figure 2.**
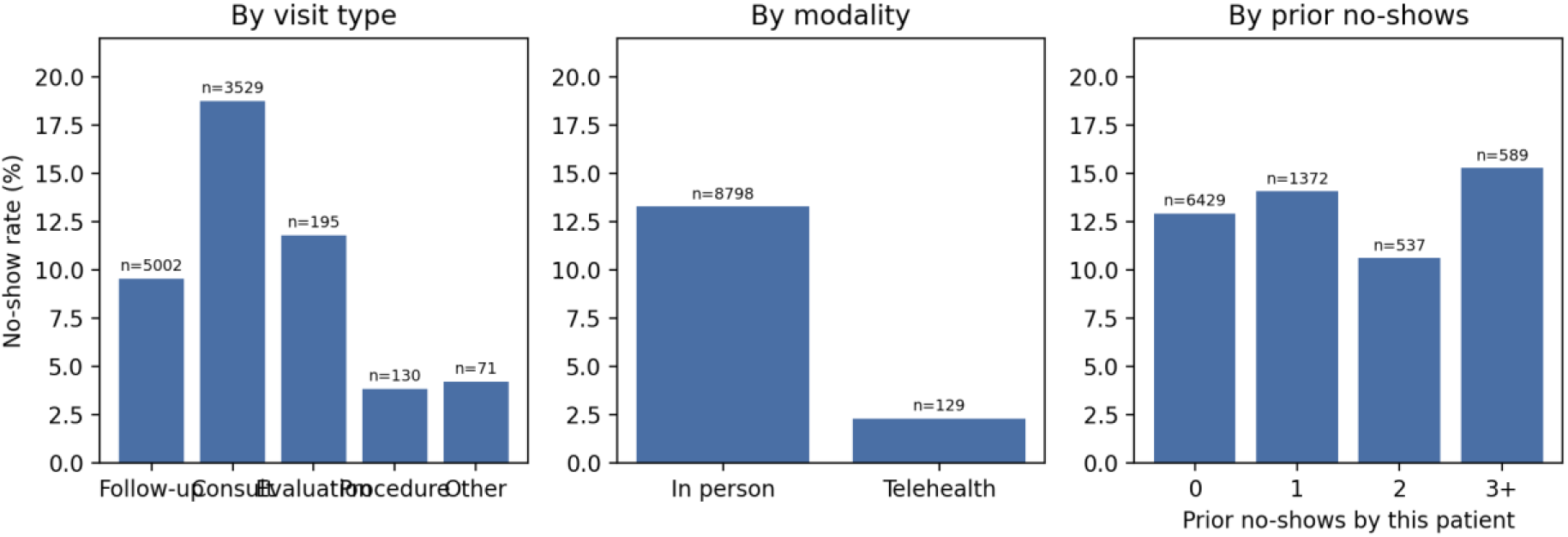
Unadjusted no-show rate by visit type, modality, and the patient’s prior no-shows in the clinic (full cohort, n = 8,927).

### Adjusted associations

In the full-cohort GEE model of scheduling predictors (Table 2), consultations were more likely than follow-ups to be missed (OR 1.25, 95% CI 1.03-1.53), while procedure visits (OR 0.36, 95% CI 0.17-0.75), telehealth visits (OR 0.39, 95% CI 0.23-0.67), and same-day appointments (OR 0.54, 95% CI 0.36-0.82) were less likely. Each prior cancellation was associated with slightly lower odds of no-show (OR 0.98, 95% CI 0.96-1.00), and the odds of no-show rose by 8% per calendar year (OR 1.08, 95% CI 1.02-1.14). Day of week, first visit, and prior no-shows were not independently associated with the outcome once the exchangeable within-patient correlation was accounted for.

**Table 2.**
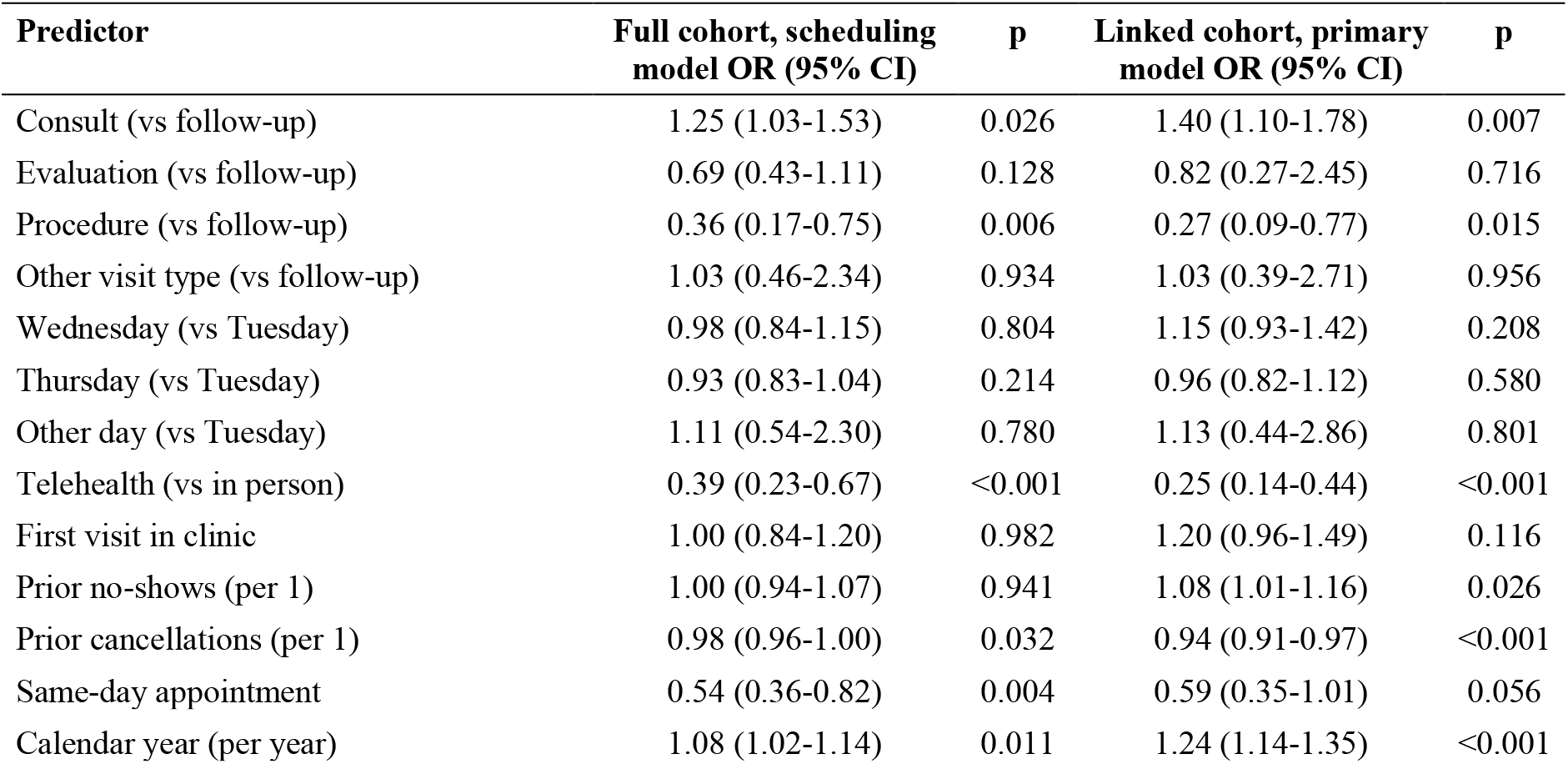
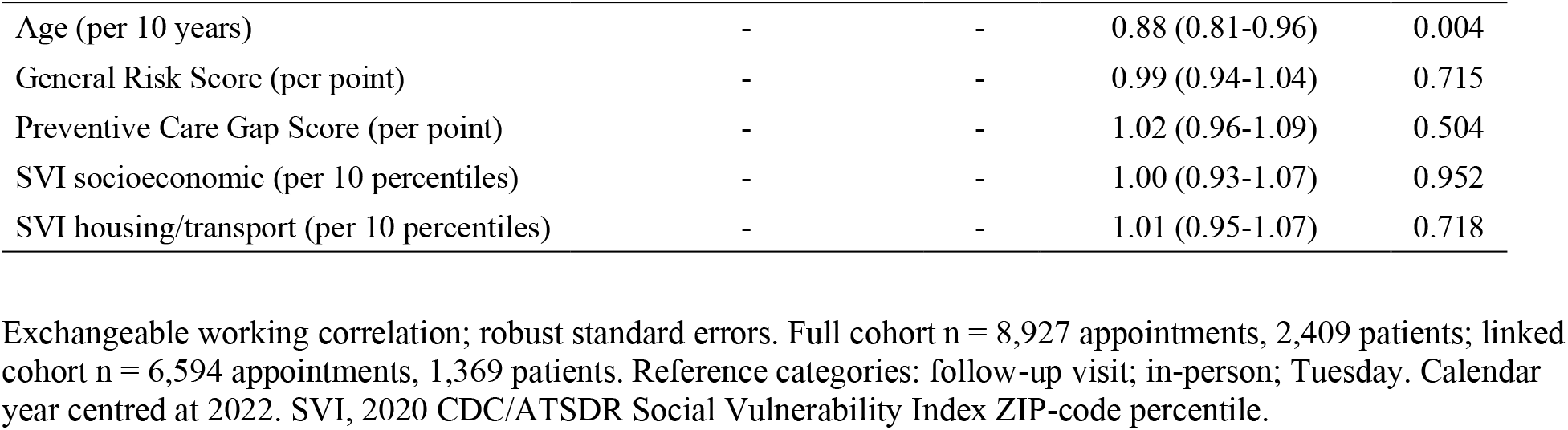
Adjusted odds of no-show per appointment (GEE, clustered by patient).

In the linked cohort (Table 2, Figure 3), the scheduling associations were similar or stronger: consultation OR 1.40, 95% CI 1.10-1.78; procedure OR 0.27, 95% CI 0.09-0.77; telehealth OR 0.25, 95% CI 0.14-0.44; same-day OR 0.59, 95% CI 0.35-1.01; prior no-shows OR 1.08, 95% CI 1.01-1.16; prior cancellations OR 0.94, 95% CI 0.91-0.97. Older age was associated with lower odds of no-show (OR 0.88, 95% CI 0.81-0.96 per decade). The EPIC General Risk Score (OR 0.99, 95% CI 0.94-1.04), Preventive Care Gap Score (OR 1.02, 95% CI 0.96-1.09), SVI socioeconomic percentile (OR 1.00, 95% CI 0.93-1.07), and SVI housing/transportation percentile (OR 1.01, 95% CI 0.95-1.07) showed no association, with confidence intervals excluding effects larger than about 6% per unit. Multicollinearity was low (maximum VIF 1.9).

**Figure 3.**
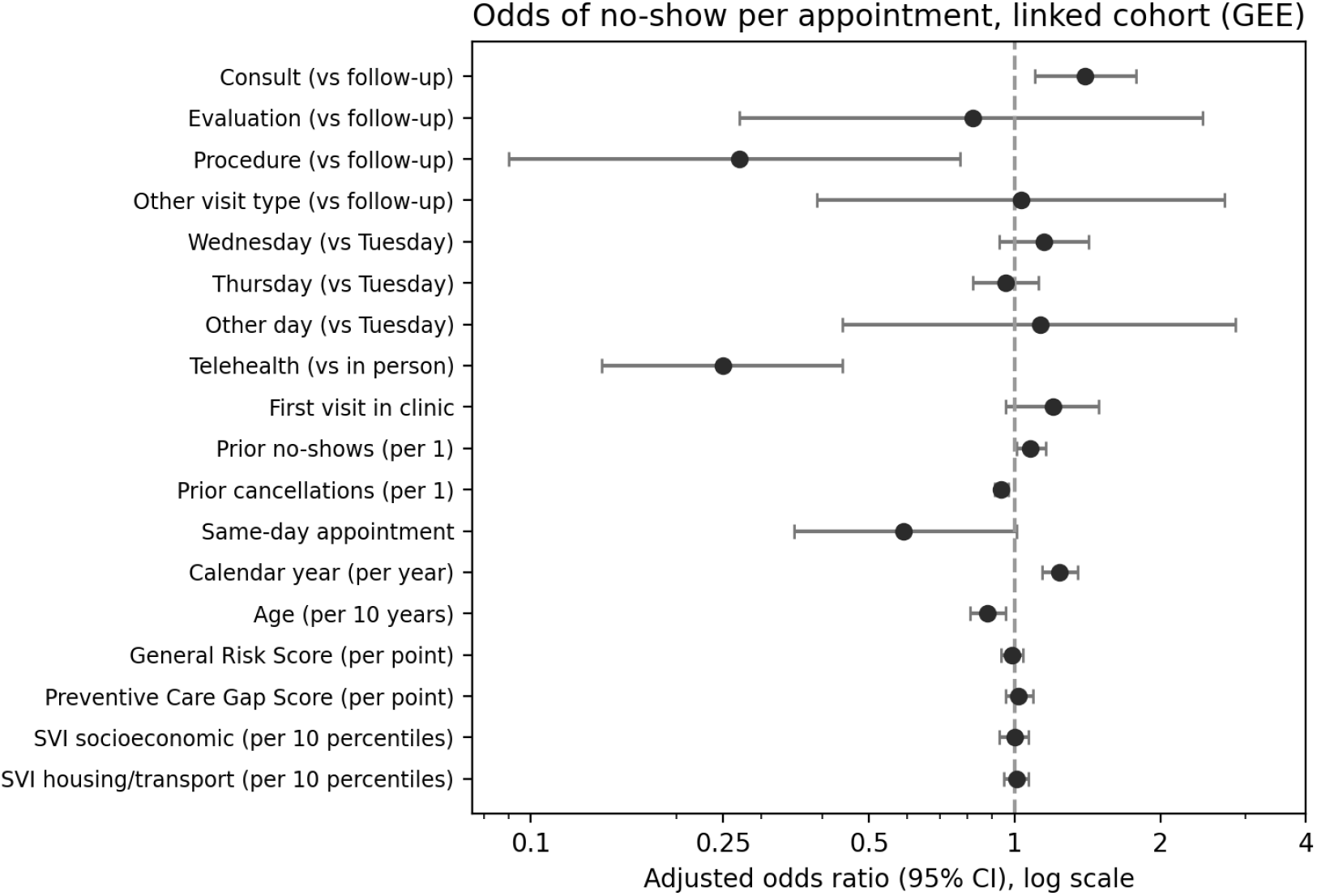
Adjusted odds ratios (95% CI) for no-show per appointment in the linked cohort (primary GEE model, Table 2).

In the fully linked cohort (Supplementary Table S1), sex (OR 0.79, 95% CI 0.60-1.04 for male), race/ethnicity (White OR 1.03, 95% CI 0.76-1.41; Black OR 0.83, 95% CI 0.56-1.23), and completion of any SDOH screening (OR 1.01, 95% CI 0.69-1.49) were not associated with no-show, and the EPIC score estimates were unchanged. In this smaller, more established subset the consultation association was not present (OR 0.75, 95% CI 0.51-1.09), consistent with its lower proportion of first visits.

### Patient-level versus appointment-level analysis

When the same 1,369 patients were analyzed at the patient level (Table 4), the General Risk Score was weakly associated with the number of PM&R no-shows (IRR 1.04 (1.00-1.09)) and, similarly, with the number of scheduled appointments (IRR 1.05 (0.99-1.11)); with scheduled appointments as an offset, the association with no-shows attenuated (IRR 1.02 (0.98-1.06)), matching the null appointment-level estimate (1.01 (0.96-1.07)). Spearman correlations of the General Risk Score with the number of appointments, the number of no-shows, and the no-show proportion were 0.07, 0.03, and 0.01. The inverse association with age was consistent across all specifications.

### Prediction

The scheduling-only logistic model in the full cohort had a patient-grouped cross-validated AUC of 0.660 (95% CI 0.636-0.681), with a calibration slope of 0.92, calibration-in-the-large of -0.00, and a scaled Brier score of 3% (Table 3, Figures 4 and 5). Gradient boosting performed similarly (AUC 0.670). Predicted risk was well ordered across deciles, from 3.4% observed no-shows in the lowest-risk decile to 22.7% in the highest (Supplementary Table S3); the highest-risk 20% of appointments contained 32% of all no-shows. In temporal validation, the model fitted to 2022-2024 appointments had an AUC of 0.624 (95% CI 0.593-0.658) on 3,085 appointments (378 no-shows) from 2025-2026, with a calibration slope of 0.60 and calibration-in-the-large of -0.18, indicating that predictions were somewhat too extreme and overestimated risk in the later period, consistent with the fall in the no-show rate after 2024.

**Figure 4.**
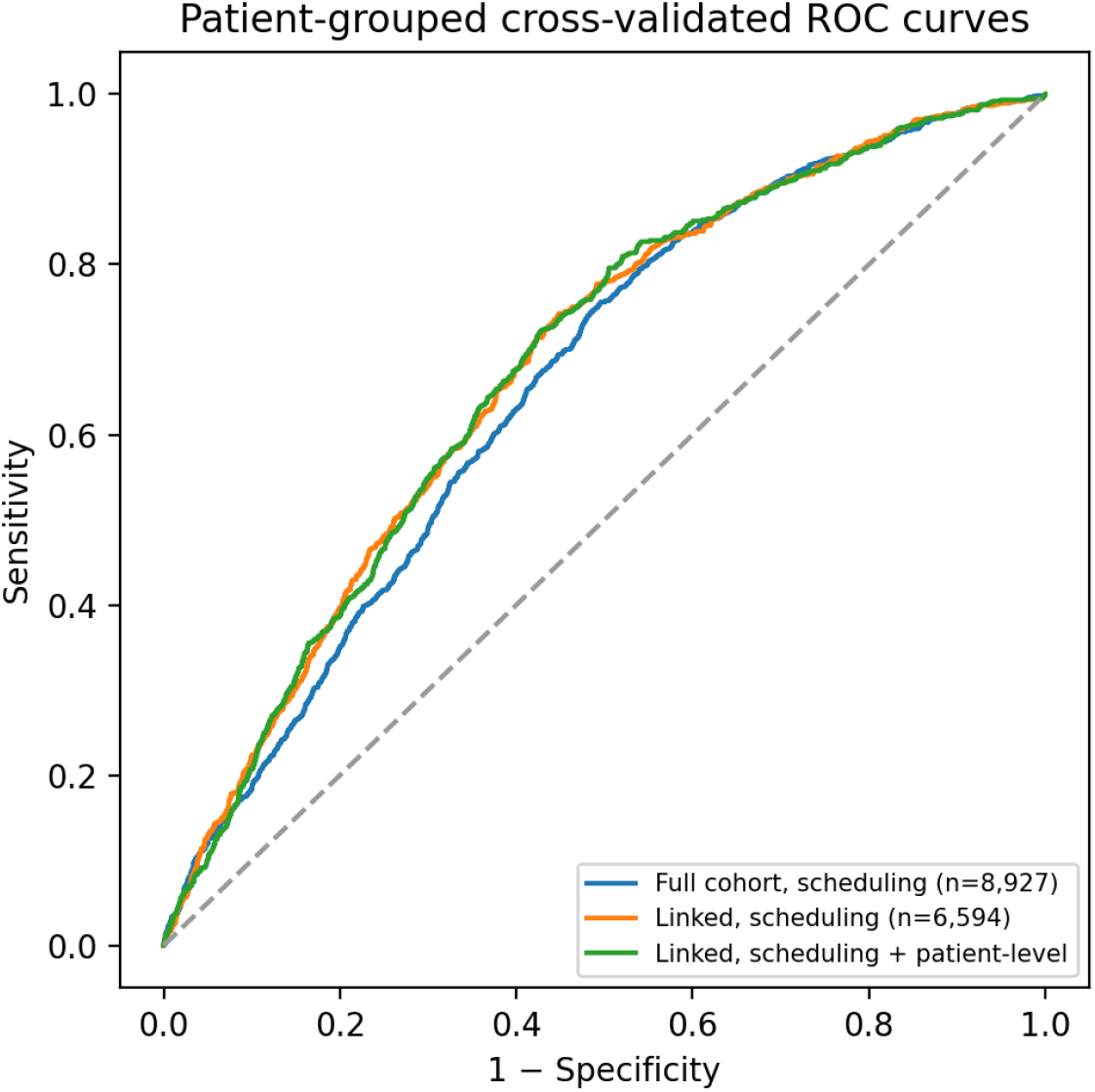
Patient-grouped cross-validated ROC curves for logistic models in the full cohort (scheduling features) and linked cohort (with and without patient-level features).

**Figure 5.**
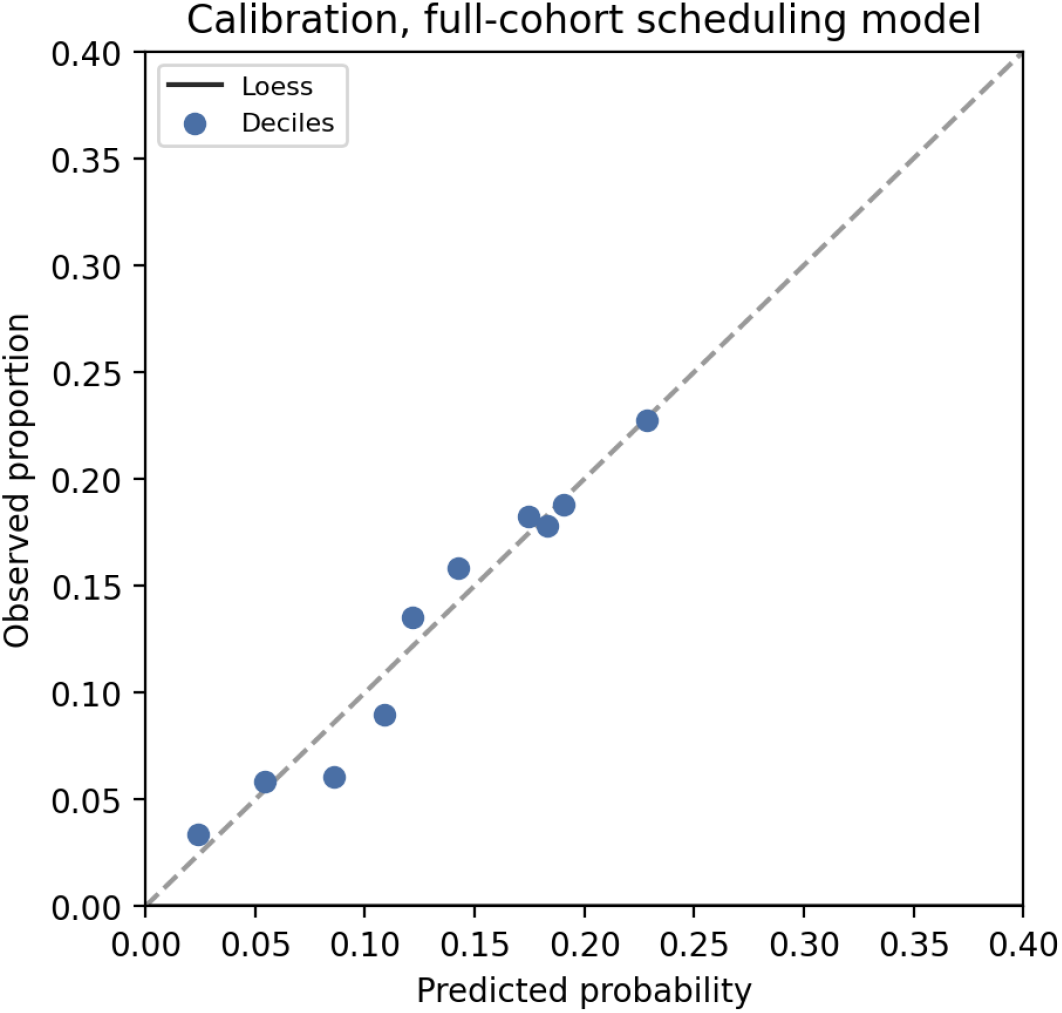
Calibration of the full-cohort scheduling model (out-of-fold predictions). Points are deciles of predicted probability; solid line, loess; dashed line, perfect calibration.

**Figure 6.**
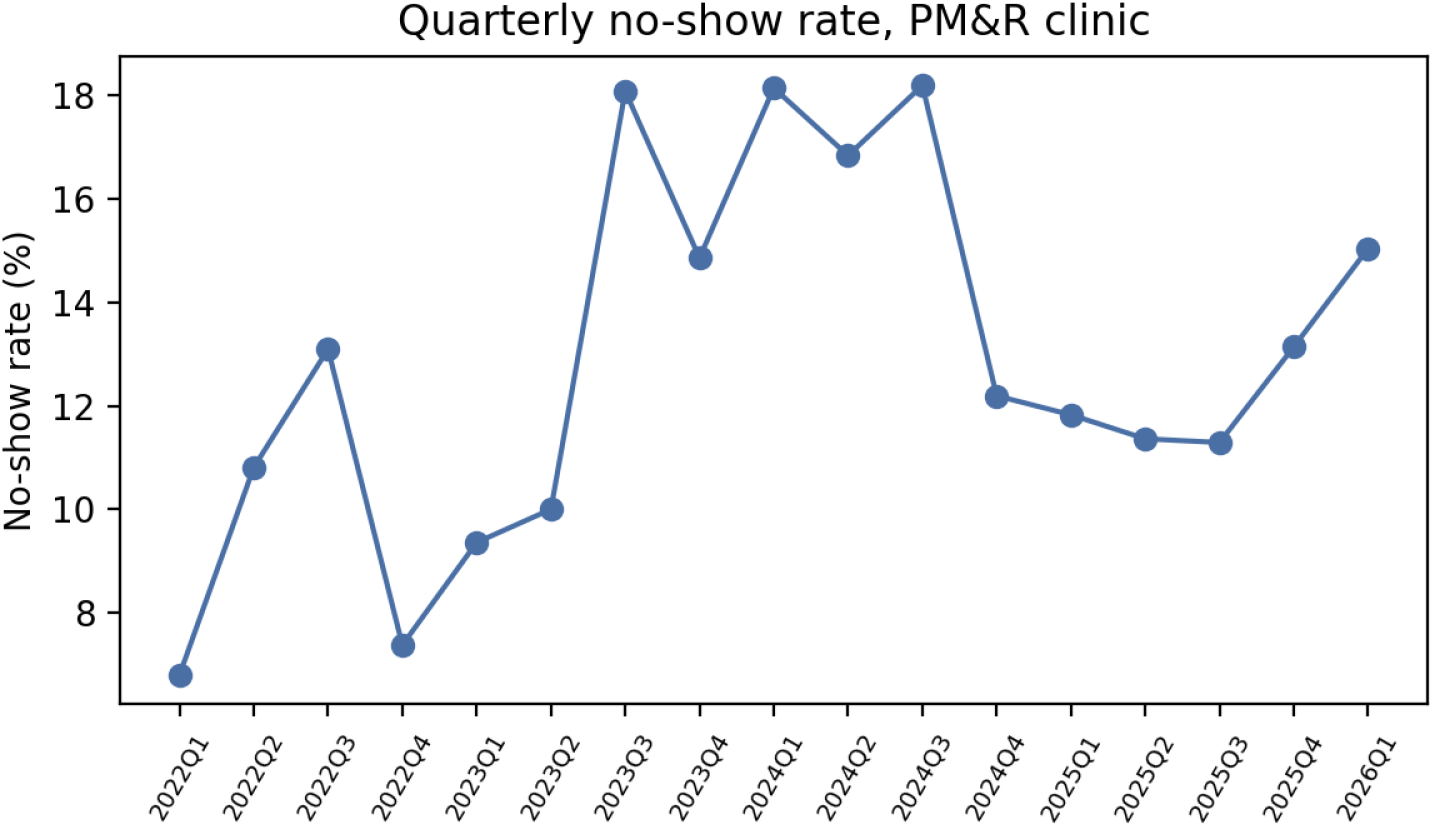
Quarterly no-show rate in the PM&R clinic, February 2022 to February 2026 (quarters with ≥50 appointments).

**Table 3.** Internal validation of no-show prediction models.

| Cohort and features | Model | Appts | Events | AUC | 95% CI | Brier | Scaled<br>Brier | Cal.<br>slope | Cal.<br>intercept |
| --- | --- | --- | --- | --- | --- | --- | --- | --- | --- |
| Full cohort, scheduling features | Logistic regression | 8,927 | 1,171 | 0.660 | 0.636-0.681 | 0.110 | 3% | 0.92 | -0.00 |
| Full cohort, scheduling features | Gradient boosting | 8,927 | 1,171 | 0.670 | 0.648-0.691 | 0.110 | 4% | 0.89 | -0.01 |
| Temporal validation: train 2022-2024, test 2025-2026 | Logistic regression | 3,085 | 378 | 0.624 | 0.593-0.658 | 0.107 | 1% | 0.60 | -0.18 |
| Linked cohort, scheduling features | Logistic regression | 6,594 | 745 | 0.679 | 0.654-0.705 | 0.096 | 4% | 0.94 | -0.00 |
| Linked cohort, scheduling features | Gradient boosting | 6,594 | 745 | 0.682 | 0.658-0.707 | 0.096 | 4% | 0.85 | -0.02 |
| Linked cohort, scheduling + patient-level features | Logistic regression | 6,594 | 745 | 0.679 | 0.653-0.703 | 0.096 | 4% | 0.92 | -0.00 |
| Linked cohort, scheduling + patient-level features | Gradient boosting | 6,594 | 745 | 0.664 | 0.634-0.692 | 0.099 | 1% | 0.74 | -0.13 |

Cross-validated rows: 5-fold cross-validation grouped by patient and stratified by outcome, repeated five times; performance on pooled out-of-fold predictions; AUC confidence intervals from 500 patient-level cluster bootstrap resamples. Temporal validation: model fitted to 2022-2024 appointments, evaluated on 2025-2026. Scaled Brier = 1 – Brier/Brier of a null model predicting the outcome prevalence. Scheduling features: visit type, telehealth, same-day, first visit, prior no-shows, prior cancellations, prior appointments, day of week, month. Patient-level features: age, General Risk Score, Preventive Care Gap Score, SVI socioeconomic and housing/transportation percentiles.

**Table 4.** Patient-level versus appointment-level estimates for the same 1,369 patients.

| Predictor | No-show count,<br>NB IRR (95% CI) | Appointment<br>count, NB IRR<br>(95% CI) | No-show count with<br>appointment offset,<br>NB IRR (95% CI) | Appointment-level GEE OR<br>(95% CI) |
| --- | --- | --- | --- | --- |
| Age (per 10 years) | 0.90 (0.83-0.98) | 1.03 (0.94-1.13) | 0.89 (0.83-0.95) | 0.86 (0.79-0.95) |
| General Risk Score (per point) | 1.04 (1.00-1.09) | 1.05 (0.99-1.11) | 1.02 (0.98-1.06) | 1.01 (0.96-1.07) |
| Preventive Care Gap Score (per point) | 1.01 (0.95-1.08) | 0.99 (0.92-1.07) | 1.03 (0.97-1.08) | 1.03 (0.97-1.10) |
| SVI socioeconomic (per 10 percentiles) | 1.00 (0.93-1.08) | 0.92 (0.84-1.01) | 1.00 (0.94-1.06) | 1.00 (0.93-1.08) |
| SVI housing/transport (per 10 percentiles) | 0.98 (0.91-1.05) | 0.98 (0.92-1.06) | 1.01 (0.96-1.06) | 1.02 (0.96-1.08) |

NB, negative binomial with HC3 robust standard errors; patient-level models include only the five predictors shown. The appointment-level column is from the patient-level-predictors-only GEE model in the linked cohort. IRR, incidence rate ratio.

In the linked cohort, adding age, EPIC scores, and SVI to the scheduling model did not improve discrimination (logistic AUC 0.679 vs 0.679; gradient boosting 0.664 vs 0.682). Discrimination was similar across age and General Risk Score strata and across sex and race/ethnicity groups, with overlapping confidence intervals (Table 6).

### Sensitivity analyses

The null associations of the EPIC scores and SVI, and the inverse association with age, were unchanged in every sensitivity analysis (Table 5). The scheduling associations were robust in direction; their magnitudes depended on the working correlation structure, with the independence structure yielding larger estimates for consultation and prior no-shows, as expected when within-patient correlation is not modeled. Telehealth could not be estimated after excluding high-volume patients because no telehealth visit in that subset was missed.

**Table 5.** Sensitivity analyses of the primary GEE model (linked cohort).

| Analysis | Appts | Consult vs<br>follow-up | Telehealth | Prior no-shows (per 1) | Age (per 10<br>y) | General Risk<br>Score |
| --- | --- | --- | --- | --- | --- | --- |
| Main model (exchangeable correlation) | 6,594 | 1.40 (1.10-1.78) | 0.25 (0.14-0.44) | 1.08 (1.01-1.16) | 0.88 (0.81-0.96) | 0.99 (0.94-1.04) |
| Independence correlation structure | 6,594 | 1.72 (1.34-2.22) | 0.17 (0.06-0.44) | 1.32 (1.24-1.40) | 0.88 (0.81-0.96) | 0.99 (0.94-1.04) |
| Excluding telehealth visits | 6,469 | 1.69 (1.31-2.18) | - | 1.32 (1.24-1.41) | 0.88 (0.81-0.96) | 0.99 (0.94-1.04) |
| Excluding same-day appointments | 6,393 | 1.45 (1.14-1.85) | 0.19 (0.09-0.41) | 1.11 (1.02-1.20) | 0.88 (0.81-0.96) | 0.99 (0.94-1.04) |
| Excluding 2026 (partial year) | 6,490 | 1.41 (1.10-1.79) | 0.24 (0.13-0.44) | 1.08 (1.01-1.17) | 0.88 (0.81-0.96) | 0.99 (0.94-1.04) |
| Excluding patients with >20 appointments | 3,471 | 1.19 (0.93-1.53) | n.e. | 1.35 (1.22-1.50) | 0.87 (0.80-0.96) | 0.99 (0.94-1.05) |

Values are OR (95% CI). Models excluding telehealth visits and patients with >20 appointments used an independence working correlation because the exchangeable model did not converge. n.e., not estimable: no telehealth visit in that subset was missed.

**Table 6.**
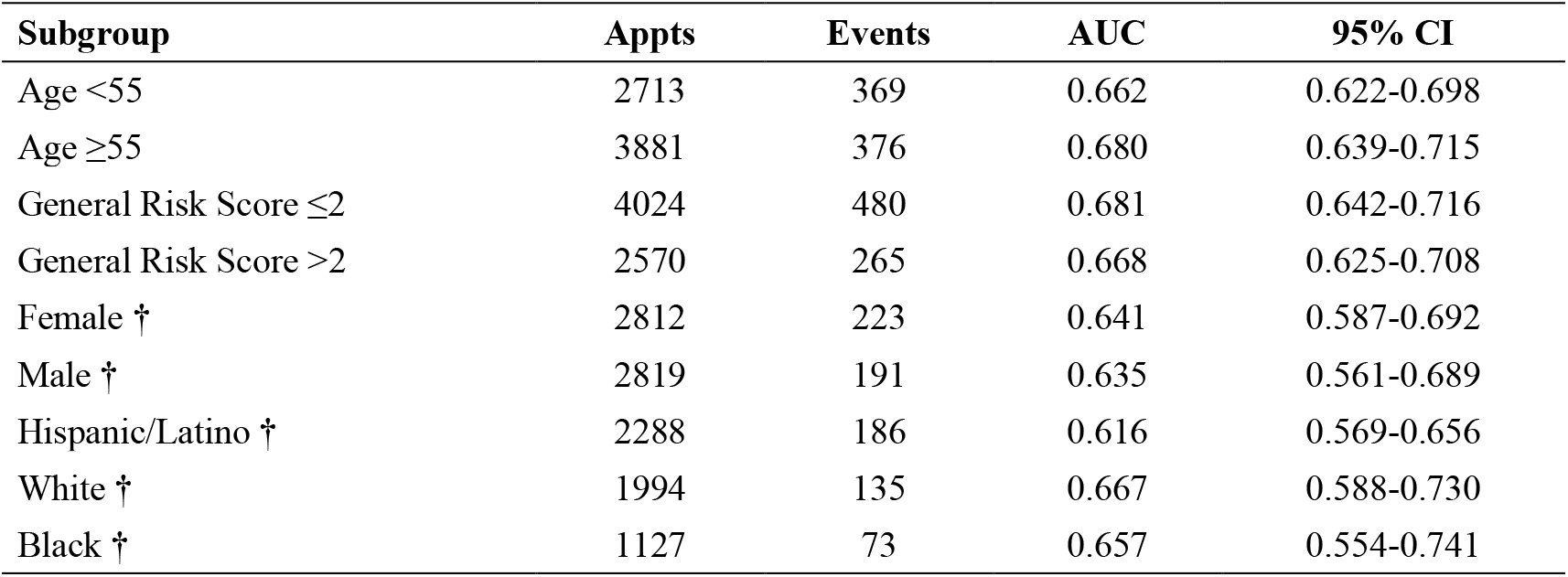
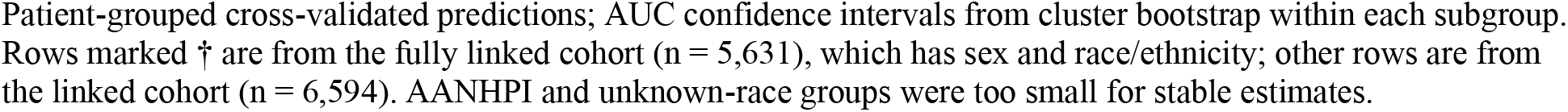
Discrimination of the scheduling + patient-level logistic model by subgroup.

| Subgroup | Appts | Events | AUC | 95% CI |
| --- | --- | --- | --- | --- |
| Age <55 | 2713 | 369 | 0.662 | 0.622-0.698 |
| Age ≥55 | 3881 | 376 | 0.680 | 0.639-0.715 |
| General Risk Score ≤2 | 4024 | 480 | 0.681 | 0.642-0.716 |
| General Risk Score >2 | 2570 | 265 | 0.668 | 0.625-0.708 |
| Female † | 2812 | 223 | 0.641 | 0.587-0.692 |
| Male † | 2819 | 191 | 0.635 | 0.561-0.689 |
| Hispanic/Latino † | 2288 | 186 | 0.616 | 0.569-0.656 |
| White † | 1994 | 135 | 0.667 | 0.588-0.730 |
| Black † | 1127 | 73 | 0.657 | 0.554-0.741 |

Patient-grouped cross-validated predictions; AUC confidence intervals from cluster bootstrap within each subgroup. Rows marked † are from the fully linked cohort (n = 5,631), which has sex and race/ethnicity; other rows are from the linked cohort (n = 6,594). AANHPI and unknown-race groups were too small for stable estimates.

## Discussion

### Principal findings

In a safety-net PM&R clinic, roughly one in eight scheduled appointments was a no-show and a further one in four was cancelled. No-shows were concentrated at new-patient consultations and in-person visits, were rare at telehealth and procedure visits, and became less likely with age.

Two composite risk scores that the EHR generates automatically, and two neighborhood social vulnerability measures, carried no information about whether an individual appointment would be missed, and adding them to a scheduling-based model did not improve prediction. A model built only from what a scheduler can see for every appointment discriminated no-shows modestly (AUC around 0.66) and identified a highest-risk fifth of appointments containing nearly a third of all no-shows.

### EHR composite scores and social vulnerability

The null result for the EPIC General Risk Score is the finding most likely to be useful to other safety-net clinics, because these scores are visible for every patient and are an obvious candidate for a no-show flag. Our comparison of analytic units shows why they may appear predictive when they are not. Patients with higher clinical risk have more appointments scheduled in the same clinic and therefore accumulate more missed visits, but they do not miss a higher proportion. An earlier patient-level analysis of this clinic, which used a missed-appointment count drawn from a hospital-wide field without a denominator, reported a strong positive association between the General Risk Score and missed appointments; the appointment-level analysis presented here does not support that conclusion. Patient-level counts without a denominator conflate utilization with non-adherence, and the effect can be large. The same logic applies to the Preventive Care Gap Score and, we suspect, to other utilization-correlated composites.

The absence of an association with ZIP-code social vulnerability is less surprising. The clinic’s catchment is concentrated in the highest-vulnerability percentiles statewide, leaving little between-patient contrast, and area-level indices are known to be weak proxies for individual social risk.4 The lack of an association with completion of SDOH screening in the fully linked cohort, in contrast to the earlier patient-level finding, likely reflects the same denominator problem: screening is completed at attended visits, so patients with more attended visits are both more likely to be screened and more likely to accumulate missed visits.

### Scheduling characteristics

The strongest associations were with characteristics of the visit itself. Telehealth visits were missed at a fraction of the rate of in-person visits, consistent with reports across specialties that virtual visits reduce no-shows by removing transportation and time costs.4,5 This is particularly relevant in a car-dependent region with limited public transit,8,12 and for a rehabilitation population with mobility limitations. Only 1.4% of appointments in this clinic were telehealth, suggesting substantial headroom. New-patient consultations were missed more often than follow-ups, consistent with weaker attachment to the clinic before a first visit, and procedure visits were rarely missed, plausibly because they are actively wanted and often preceded by preparation. Same-day appointments were also rarely missed, which supports the use of same-day slots for backfill. Prior cancellations were weakly protective, suggesting that patients who cancel are engaged enough to communicate; prior no-shows were modestly predictive in the linked cohort, less so than in other settings,5,7 perhaps because the clinic’s retention policy means that high-frequency non-attenders remain scheduled regardless of history.

The inverse association with age was robust across all specifications, in contrast to the earlier patient-level analysis where it depended on a few extreme patients. Older patients in this clinic, many of whom are within Medi-Cal, Workers’ Compensation, or disability programs whose continuation depends on documented follow-up, may face structural incentives to attend that outweigh mobility barriers. This remains a hypothesis; the data do not identify the mechanism.

The rise in no-shows from 2022 to 2024 and fall in 2025 was not explained by measured predictors and degraded temporal validation, with the model overestimating risk in 2025-2026. Clinic staffing, scheduling practices, or reminder systems may have changed over the period; we could not measure these. Any deployed model would need periodic recalibration.

### Comparison with prior work

Machine-learning no-show models in radiology, primary care, and breast imaging have reported AUCs from roughly 0.70 to 0.85, generally by drawing on appointment lead time, prior no-show history, and reminder-response data.5,6,15 Our discrimination is at the lower end of this range, most likely because booking date, and therefore lead time-consistently among the strongest predictors-was not available, and because the clinic’s retention policy weakens the usual signal from prior no-shows. That a flexible learner added nothing over logistic regression is consistent with systematic reviews finding that feature quality, not model complexity, determines performance in this domain.7 Our contribution is less the model than the negative result for EHR composites and social indices, and the demonstration that unit of analysis can reverse conclusions.

### Use in practice

The scheduling-only model uses six fields available in any Epic scheduling view-visit type, modality, same-day status, calendar period, and the patient’s counts of prior no-shows and cancellations in the clinic-and its equation is provided in Supplementary Table S2. No patient-level clinical data are required, so it can be applied at booking. The output is a probability; Supplementary Table S3 shows observed no-show rates by predicted-risk decile so that a clinic can select a threshold matched to its outreach capacity. Given the modest discrimination, the model is suited to prioritizing reminder calls or offering telehealth conversion, not to overbooking decisions. It has not been externally validated, and calibration drifted within the study period, so performance should be monitored if deployed.

### Limitations

This study is retrospective and single-centre; associations are not causal, and the clinic’s three-day schedule, retention policy, and regional characteristics may limit generalizability. Booking date was unavailable, so lead time could not be modeled, and reminder-call data were not captured. Patient-level predictors were taken at the time of extraction rather than at each appointment; for the EPIC scores this means a 2025 value was attached to appointments as early as 2022, which biases associations toward the null only if scores drift randomly, and toward spurious association if scores rise with utilization. That we found no association makes the second concern moot for our conclusions, but a prospective study should use point-in-time scores. Age was likewise measured at extraction. Patient-level data were available for 57% of patients, and those without it were newer to the clinic with a higher no-show rate; the primary adjusted model therefore under-represents first visits, which is why the scheduling-only model was fitted in the full cohort. Sex, race/ethnicity, and SDOH screening were available for only 37% of patients, in a subset with an unusually low no-show rate, limiting power to detect demographic differences. The no-show rate changed over time for reasons we could not measure. Finally, the earlier preprint from this clinic reported different conclusions from a patient-level analysis; we have set out above why the appointment-level analysis should be preferred.

### Future directions

Obtaining booking dates and reminder-system logs would likely improve prediction substantially. Point-in-time EPIC scores would allow a cleaner test of whether clinical risk affects attendance. The strong telehealth association warrants a pragmatic trial of offering telehealth conversion to appointments flagged as high risk. And structured interviews with high-frequency non-attenders, potentially analyzed with natural language processing,13 could identify barriers that structured scheduling data cannot.

## Conclusion

In a safety-net PM&R clinic, no-shows were associated with the characteristics of the visit-new-patient consultations, in-person modality, and advance rather than same-day scheduling-and with younger age, but not with EHR composite risk scores, neighborhood social vulnerability, sex, race/ethnicity, or SDOH screening completion. Composite risk scores that appear predictive in patient-level counts do not predict non-attendance at the appointment level. Efforts to reduce missed visits in similar settings should focus on scheduling-level levers, particularly telehealth access and outreach before first visits, rather than on clinical risk stratification.

## Supplementary material

**Supplementary File 1.** Analysis code.

**Supplementary File 2.** TRIPOD+AI checklist.

## Supporting information

Supplementary File 2. TRIPOD+AI checklist

Supplementary File 1. Analysis code (Python)

## Data Availability

Data availability. Appointment-level data cannot be shared publicly because they are governed by the hospital's data-use policies. De-identified aggregate data supporting the tables are available from the corresponding author for non-commercial research purposes, subject to institutional approval. A data dictionary is provided in Supplementary Table S4.
Code availability. The complete analysis code (Python), which reproduces every number, table, and figure from the analytic dataset, is provided as Supplementary File 1.

---

**Supplementary Table S1.** GEE model in the fully linked cohort (n = 5,631 appointments, 887 patients), adding sex, race/ethnicity, and SDOH screening.

| Predictor | OR | 95% CI | p |
| --- | --- | --- | --- |
| Consult (vs follow-up) | 0.75 | 0.51-1.09 | 0.131 |
| Evaluation (vs follow-up) | 0.84 | 0.18-3.92 | 0.821 |
| Procedure (vs follow-up) | 0.35 | 0.11-1.12 | 0.077 |
| Other visit type (vs follow-up) | 1.28 | 0.49-3.39 | 0.614 |
| Wednesday (vs Tuesday) | 1.10 | 0.82-1.47 | 0.531 |
| Thursday (vs Tuesday) | 0.97 | 0.80-1.19 | 0.789 |
| Other day (vs Tuesday) | 0.81 | 0.21-3.09 | 0.761 |
| Male sex | 0.79 | 0.60-1.04 | 0.089 |
| White (vs Hispanic/Latino) | 1.03 | 0.76-1.41 | 0.838 |
| Black (vs Hispanic/Latino) | 0.83 | 0.56-1.23 | 0.343 |
| AANHPI (vs Hispanic/Latino) | 0.70 | 0.23-2.15 | 0.531 |
| Unknown race (vs Hispanic/Latino) | 1.60 | 0.87-2.96 | 0.129 |
| Telehealth (vs in person) | 0.24 | 0.12-0.48 | <0.001 |
| First visit in clinic | 0.80 | 0.53-1.21 | 0.296 |
| Prior no-shows (per 1) | 1.08 | 0.99-1.19 | 0.092 |
| Prior cancellations (per 1) | 0.96 | 0.93-1.00 | 0.046 |
| Same-day appointment | 0.66 | 0.32-1.35 | 0.253 |
| Calendar year (per year) | 1.14 | 1.01-1.28 | 0.031 |
| Age (per 10 years) | 0.84 | 0.75-0.95 | 0.007 |
| General Risk Score (per point) | 1.03 | 0.95-1.11 | 0.472 |
| Preventive Care Gap Score (per point) | 1.03 | 0.92-1.15 | 0.586 |
| SVI socioeconomic (per 10 percentiles) | 1.08 | 0.97-1.21 | 0.176 |
| SVI housing/transport (per 10 percentiles) | 1.00 | 0.92-1.09 | 0.945 |
| Any SDOH screening recorded | 1.01 | 0.69-1.49 | 0.946 |

**Supplementary Table S2.** Full-cohort scheduling-only logistic regression equation (log-odds coefficients, unstandardized). Predicted probability = 1 / (1 + exp(−(intercept + Σ coefficient × value))).

| Term | Coefficient (SE) |
| --- | --- |
| Intercept | -1.3538 (0.8031) |
| Telehealth | -1.7847 (0.6666) |
| First visit | 0.0688 (0.0847) |
| Prior no-shows | 0.2845 (0.0268) |
| Prior cancellations | 0.0612 (0.0243) |
| Prior appointments | -0.0590 (0.0084) |
| Same-day | -0.6107 (0.2340) |
| Month (1-12) | -0.0033 (0.0093) |
| Visit type: CONSULT | 0.3379 (0.0899) |
| Visit type: EVALUATION | -0.2284 (0.2399) |
| Visit type: PROCEDURE | -1.1327 (0.4624) |
| Visit type: OTHER | 0.2349 (0.6948) |
| Day of week: 1 | -0.4848 (0.7998) |
| Day of week: 2 | -0.4368 (0.8011) |
| Day of week: 3 | -0.5524 (0.7999) |
| Day of week: 4 | -0.7456 (1.0058) |

Fitted to all 8,927 appointments. Day of week coded as Python weekday (1 = Tuesday, 2 = Wednesday, 3 = Thursday). Indicator terms coded 1 = present.

**Supplementary Table S3.**
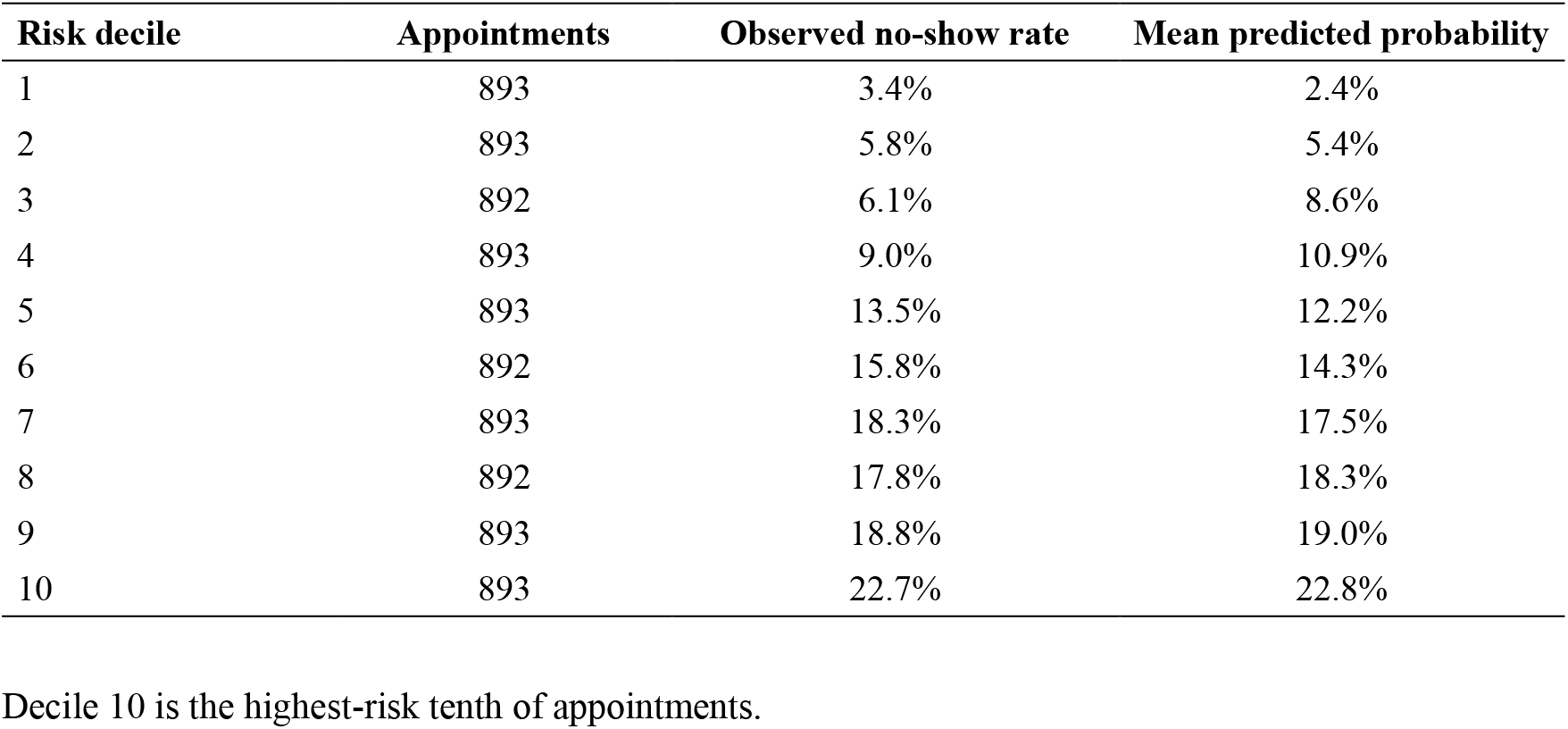
Observed and predicted no-show rate by decile of predicted risk, full-cohort scheduling model (out-of-fold).

**Supplementary Table S4.** Data dictionary.

| Variable | Source | Definition / coding |
| --- | --- | --- |
| noshow | Scheduling extract | Appointment Status = “No Show”; 1/0. “Canceled” and “Completed” coded 0. |
| vtype | Scheduling extract | Visit Type collapsed to Follow-up / Consult / Evaluation / Procedure / Other. |
| tele | Scheduling extract | Telehealth flag present; 1/0. |
| sameday | Scheduling extract | Same-day appointment flag; 1/0. |
| dow, month, year | Scheduling extract | Derived from appointment date. |
| first_visit, prior_n, prior_ns, prior_cx | Derived | From the patient's earlier-dated PM&R appointments in the study window only. |
| age | Risk-score extract | Age in years at extraction. |
| grs, pcg | Risk-score extract | EPIC General Risk Score; EPIC Preventive Care Gap Score; values at extraction. |
| svi_ses, svi_hous | Risk-score extract | CDC/ATSDR 2020 SVI ZIP-code percentile: socioeconomic; housing type & transportation. |
| sex, race | Demographic extract | Legal sex (0 female, 1 male); EHR race/ethnicity field. |
| any_reported | Demographic extract | 1 if any of 12 EPIC SDOH domains has a recorded result. |

## Notes

### Competing Interest Statement

The authors have declared no competing interest.

### Author Declarations

Ethics committee/IRB of Arrowhead Regional Medical Center gave ethical approval for this work.

### Summary of Updates

Analysis rewritten at the appointment level. Version 1 analyzed patient-level counts of missed appointments drawn from a hospital-wide field, without a denominator of scheduled visits, and used cross-validation split by record rather than by patient, which inflated performance estimates. Version 2 uses one row per scheduled PM&R appointment (8,927 appointments, 2,409 patients, February 2022 to February 2026), generalized estimating equations clustered by patient, and patient-grouped and temporal validation. Study period corrected to reflect Epic scheduling data availability. Conclusions changed: EHR composite risk scores and neighborhood social vulnerability are not associated with no-show at the appointment level; visit characteristics and age are. Title, abstract, methods, results, tables, and figures rewritten. Analysis code and TRIPOD+AI checklist added.

