## Supplementary File 2. TRIPOD+AI checklist for "Prediction Modeling of Missed Appointments in Safety-Net Physiatry: Visit Characteristics Outperform Patient Risk Scores"

**Supplementary File 2. TRIPOD+AI checklist for the reporting of prediction model studies**

Manuscript: Appointment-Level Predictors of Missed Visits in Safety-Net Physiatry (version 2). Checklist from Collins GS et al., BMJ 2024;385:e078378. Section references are to the manuscript; replace with page numbers at submission. Items marked "AUTHORS TO COMPLETE" require information not available to the analyst.

| **Section/Topic** | **Item** | **Checklist item** | **Reported in (section)** |
| --- | --- | --- | --- |
| **TITLE** | | | |
| Title | 1 | Identify the study as developing or evaluating a multivariable prediction model, the target population, and the outcome | Title |
| **ABSTRACT** | | | |
| Abstract | 2 | See TRIPOD+AI for Abstracts checklist | Abstract (structured) |
| **INTRODUCTION** | | | |
| Background | 3a | Healthcare context, rationale, references to existing models | Introduction ¶1–2 |
|  | 3b | Target population, intended purpose in the care pathway, intended users | Introduction ¶4 (aim 3) |
|  | 3c | Known health inequalities between sociodemographic groups | Introduction ¶3 |
| Objectives | 4 | Study objectives; development and/or validation | Introduction ¶4 |
| **METHODS** | | | |
| Data | 5a | Data sources, rationale, representativeness | Methods: Study design and setting; Participants |
|  | 5b | Dates of data collection / accrual / follow-up | Methods: Study design and setting |
| Participants | 6a | Setting, number and location of centres | Methods: Study design and setting |
|  | 6b | Eligibility criteria | Methods: Participants |
|  | 6c | Treatments received, if relevant | Not applicable (no treatment variable) |
| Data preparation | 7 | Pre-processing and quality checks; similar across groups | Methods: Data sources and linkage; Suppl. File 1 |
| Outcome | 8a | Outcome definition, time horizon, assessment, consistency across groups | Methods: Outcome |
|  | 8b | Qualifications of outcome assessors if subjective | Not applicable (administrative record) |
|  | 8c | Blinding of outcome assessment | Methods: Outcome |
| Predictors | 9a | Choice and pre-selection of predictors | Methods: Predictors (final ¶) |
|  | 9b | Definition and timing of all predictors; blinding | Methods: Predictors; Suppl. Table S2 |
|  | 9c | Qualifications of predictor assessors if subjective | Not applicable |
| Sample size | 10 | How study size was arrived at; justification | Methods: Statistical analysis (Sample size) |
| Missing data | 11 | Handling of missing data; reasons for omitting data | Methods: Participants; Results ¶1 |
| Analytical methods | 12a | How data were used; partitioning; leakage | Methods: Statistical analysis (patient-grouped CV; temporal validation) |
|  | 12b | Handling of predictors (functional form, scaling) | Methods: Statistical analysis |
|  | 12c | Model type, rationale, model-building, tuning, internal validation | Methods: Statistical analysis |
|  | 12d | Heterogeneity across clusters | Not applicable (single centre) |
|  | 12e | Performance measures and plots | Methods: Statistical analysis (AUC, calibration, Brier, subgroup) |
|  | 12f | Model updating | Not applicable (no updating) |
|  | 12g | How predictions were calculated (evaluation) | Suppl. Table S1; Suppl. File 1 |
| Class imbalance | 13 | Class-imbalance methods | Methods: Statistical analysis (none used) |
| Fairness | 14 | Approaches to model fairness | Methods: Statistical analysis; Results: Table 6 |
| Model output | 15 | Output type; thresholds and rationale | Methods: Statistical analysis; Discussion: Use of the model |
| Training vs evaluation | 16 | Differences between development and evaluation data | Methods: Statistical analysis (temporal validation period stated) |
| Ethical approval | 17 | IRB name; consent or waiver | Declarations (ARMC IRB #24-29) |
| **OPEN SCIENCE** | | | |
| Funding | 18a | Funding source and role | Declarations — AUTHORS TO COMPLETE |
| Conflicts of interest | 18b | Conflicts for all authors | Declarations — AUTHORS TO COMPLETE |
| Protocol | 18c | Protocol availability | Declarations |
| Registration | 18d | Registration information | Declarations |
| Data sharing | 18e | Data availability | Declarations |
| Code sharing | 18f | Code availability | Declarations; Suppl. File 1 |
| **PATIENT & PUBLIC INVOLVEMENT** | | | |
| PPI | 19 | Patient and public involvement | Declarations |
| **RESULTS** | | | |
| Participants | 20a | Flow of participants; outcome counts; diagram | Results ¶1; Figure 1 |
|  | 20b | Characteristics; sample size; events; missing data | Results: Table 1 |
|  | 20c | Comparison of evaluation vs development data | Results: Prediction (temporal validation); Methods: Data sources (linked vs unlinked) |
| Model development | 21 | Participants and events in each analysis | Results ¶1; Figure 1 |
| Model specification | 22 | Full model to allow predictions in new individuals | Suppl. Table S2 |
| Model performance | 23a | Performance with CIs, including subgroups; plots | Results: Table 3, Table 6, Figures 4 and 5 |
|  | 23b | Heterogeneity across clusters | Not applicable |
| Model updating | 24 | Results of model updating | Not applicable |
| **DISCUSSION** | | | |
| Interpretation | 25 | Overall interpretation incl. fairness, in context of prior studies | Discussion: Principal findings; Interpretation; Comparison with prior work |
| Limitations | 26 | Limitations and their effects | Discussion: Limitations |
| Usability | 27a | Handling of poor-quality or unavailable input data | Discussion: Use of the model in practice |
|  | 27b | User interaction and expertise required | Discussion: Use of the model in practice |
|  | 27c | Next steps for research; applicability and generalizability | Discussion: Future directions |
